# Vaccine Rollout Strategies: The Case for Vaccinating Essential Workers Early

**DOI:** 10.1101/2021.02.23.21252309

**Authors:** N. Mulberry, P. Tupper, E. Kirwin, C. McCabe, C. Colijn

**Affiliations:** Department of Mathematics, Simon Fraser University; Institute of Health Economics, Alberta, Canada; Health Organisation, Policy, and Economics, School of Health Sciences, University of Manchester, United Kingdom; Department of Emergency Medicine, Faculty of Medicine and Dentistry, University of Alberta, Canada

## Abstract

In planning for upcoming mass vaccinations against COVID-19, many jurisdictions have proposed using primarily age-based rollout strategies, where the oldest are vaccinated first and the youngest last. In the wake of growing evidence that approved vaccines are effective at preventing not only adverse outcomes, but also infection (and hence transmission of SARS-CoV-2), we propose that such age-based rollouts are both less equitable and less effective than strategies that prioritize essential workers. We demonstrate that strategies that target essential workers earlier consistently outperform those that do not, and that prioritizing essential workers provides a significant level of indirect protection for older adults. This conclusion holds across numerous outcomes, including cases, hospitalizations, Long COVID, deaths and net monetary benefit, and over a range of possible values for the efficacy of vaccination against infection. Our analysis focuses on regimes where the pandemic continues to be controlled with distancing and other measures as vaccination proceeds, and where the vaccination strategy is expected to last for over the coming 6-8 months — for example British Columbia, Canada. In such a setting with a total population of 5M, vaccinating essential workers sooner is expected to prevent over 200,000 infections, over 600 deaths, and to produce a net monetary benefit of over $500M.

## 1 Introduction

Since the novel coronavirus SARS-CoV-2 emerged in late 2019, over 100 million cases of COVID-19 have been reported worldwide, resulting in over 2.4 million deaths^1^. For the first year of the pandemic, the main tools in controlling the virus’ spread have been non-pharmaceutical interventions (NPI), such as lockdowns, social distancing and personal protective equipment including face masks. But starting in late 2020, vaccines that had been in development since the start of the pandemic finished their Phase 3 trials and began to be approved in some jurisdictions. As of February 2021, four vaccines have received widespread approval for at least emergency use^62^.

Vaccinations have begun in many parts of the world, but the quantity of vaccine doses available differs greatly between jurisdictions. Shortages of vaccines in many places mean it will be many months before everyone who wants a vaccine receives one, and so decisions have to be made about who receives vaccines first. The question of how to deploy vaccines, taking into account their efficacy in preventing symptomatic disease, their efficacy in blocking transmission, the underlying contact structure of the population, the evolution of the virus, and changing non-pharmaceutical measures, poses substantial modelling challenges. An important early study was Bubar et al^15^ who modeled the pandemic using an age-stratified SEIR model and compared five vaccine prioritization strategies. They found that targeting 20–49-year-olds reduced the overall number of infections, but led to higher mortality among the elderly. Another approach is that of Chen et al^20^ who used an agent-based model with a detailed social contact network to study vaccine prioritization. They found that targeting individuals with many contacts rather than a purely age-based strategy lead to substantially better outcomes. Other studies are also split, and favour vaccinating essential workers early under some circumstances^16, 33^ or a primarily age-based prioritization in others^10^.

A natural goal for minimizing the impact of the pandemic is to prevent as many deaths due to COVID-19 as possible. This is a primary motivation for vaccination plans that start with the oldest individuals and then go down through the age cohorts, since risk of mortality increases sharply with age^39^. But another important consideration is that, in a small but significant number of cases at all ages, COVID causes long-lasting symptoms that can be debilitating^61^. There is a syndrome that has come to be known as *Long COVID*^24^: extreme fatigue and other COVID symptoms that may last for weeks or months^38^, and may turn out to be chronic to the best of our knowledge now^61^. The symptoms are similar to those described by survivors of SARS^35^, and fit the clinical definition of myalgic encephalomyelitis/chronic fatigue syndrome (ME/CFS)^27^. In addition, there are *COVID complications*^28^: severe secondary conditions caused by COVID-19 infection, including diabetes^51^, organ damage^45, 50^, and neurological and psychiatric disorders^56^. These can also be caused by other severe viral infections such as SARS^31, 35^ and Zika virus^18, 52^, and early evidence suggests that they are not rare for hospitalized COVID cases. Both of these outcomes, Long COVID and COVID complications, which we will together refer to as *chronic outcomes*, are likely to be lasting and serious consequences of the pandemic for many individuals^53^. Accordingly, vaccination strategies need to factor them into consideration, despite uncertainty in the duration, severity, and frequency of their occurrence.

Another aspect of the pandemic is that not all communities are affected equally. Disadvantaged racial and socioeconomic groups have been more severely hit by the pandemic, with higher rates of infection and death^21, 47, 49^. One proximal factor partially explaining this difference is that such groups are disproportionately represented among essential workers^19^, whom we define as workers whose employment requires them to have many contacts outside of the home. For this reason, the timing of vaccination for essential workers will have consequences not only for transmission and exposure in the population, but for equity.

Recent phase 3 trials have shown that the Moderna vaccine^11^, the Pfizer-BioNTech vaccine^43^, and the AstraZeneca^57^ vaccine are effective at preventing symptomatic infection and severe illness. There are broadly two ways that a vaccine can attain this kind of result: either by preventing infection from occurring in the first place (“sterilizing immunity”) or allowing infection but preventing disease^17^. In the first case the vaccine necessarily prevents onward transmission, but in the latter case the vaccine may or may not prevent subsequent transmission, depending in part on whether it decreases viral load. The first published reports of Phase 3 trials did not directly address this question, only reporting on the reduction of symptomatic and severe infection. However, knowledge of how effective vaccines are at preventing transmission is crucial for determining optimal vaccination rollout strategies. The emerging data relevant to this issue show both a high rate of sterilizing immunity and a reduction of viral load in the minority of those vaccinated who do become infected. One preliminary set of such data was obtained during Phase 3 trials for the Moderna vaccine: subjects received a PCR test with their second dose, thereby allowing an estimate of the efficacy of the first dose in preventing infection. A 2/3 reduction in infection was observed in *asymptomatic* infection^42^, which together with the already documented reduction in symptomatic cases^11^ gives a high overall reduction in infection. Similar preliminary results are emerging for the Pfizer vaccine^32, 41^ and for AstraZeneca vaccine^6^. As well, substantial reduction in viral loads have been found among those who are infected for both the AstraZeneca vaccine^25^ and the BioNTech/Pfizer BNT162b2 vaccine^37, 44^, suggesting reduced further transmission even when infection does occur. Given these data, the impact of vaccination on transmission is likely to be strong, and jurisdictions which have planned predominantly oldest-first vaccination rollouts may now wish to use vaccination to reduce transmission more rapidly.

Building on the work of Bubar et al^15^, we developed an age- and contact-structured model to explore the impact of vaccination strategies. We incorporate chronic outcomes of COVID-19 infection, along with infections, hospitalizations and deaths. We compare age-based vaccine rollout from oldest to youngest with strategies that prioritize those we call “essential workers” – those whose employment requires them to have considerable contact outside the home. Furthermore, we attach health economic outputs by applying a Net Benefit framework to our results, including the expected cost due to hospitalization and chronic conditions resulting from infection, as well as decrements in health utility, which are measured using quality-adjusted life years (QALYs). These estimates are synthesized into Net Monetary Benefit (NMB) by converting QALYs to monetary value, to allow a unified way of evaluating vaccination strategies^54^.

## 2 Material and Methods

We use an age-structured compartmental model to investigate the impact of different vaccination strategies in British Columbia, following Bubar et al^15^. The model has susceptible, exposed, infectious and recovered individuals and was originally developed to explore vaccination by age, considering “leaky” or “all or nothing” vaccination and taking existing seroprevalence into account. Our focus is different; we extend the model by considering groups separated by both age and essential worker status, and we explore Long COVID and chronic outcomes. We also have two distinct efficacy modes: preventing infection and therefore transmission, and preventing severe outcomes (symptomatic disease, hospitalization, death and chronic outcomes). We have made the code necessary ot reproduce our results publicly available ^1^.

In total, we model groups {0*−* 9, 10*−* 19, 20 *−*29, …, 70 *−*79, 80+, 20*−*29^*e*^, 30*−*39^*e*^, …, 70*−*79^*e*^}, where the *e* superscript denotes an “essential worker” group. We furthermore extend Bubar et al’s analysis by implementing and simulating more detailed age-based and essential-worker-based vaccination strategies, and by considering trade-offs between the efficacy of vaccination on preventing infection, and on preventing adverse outcomes.

To model contacts among all 15 groups, we first start with the four matrices Λ_*R*_ developed by Prem et al^46^ for estimated contact patterns among Canadians at home, work, school, and all other locations. We assume that the average contact rates between each age group and at each location are the same across the population. Each contact matrix Λ_*R*_ provides an estimate of contact patterns over age groups 0–70+ in five-year increments. We first combine each age group into the desired ten-year bins using demographic data from British Columbia (BC), Canada. We then obtained survey data to estimate the distribution of essential workers by age group (the Covid Speak Survey^3^); these proportions varied from 20% of 30–39 years-olds to approximately 10% of 70–79 years-olds. In total, we estimate that approximately 13% of the population of BC are considered to be “essential”. We use this data to split the working adult age groups into essential worker and non-essential worker compartments, assuming an even distribution of the number of contacts, following the technique of Buckner et al^16^. We model social distancing in part by eliminating workplace contact between non-essential workers (schools have remained open in BC, and so we do not eliminate school-based contact). In addition, all other contacts are then reduced, reflecting broad distancing measures. By modifying the scaling factor we control the reproductive number, *R*. In our formulation *R* reflects the extent and effectiveness of NPI measures (but does not reflect the impact of vaccination). The underlying transmission (parameterized by *R*) impacts the relative benefits of different vaccination strategies, and we explore this impact.

Our simulation approach is motivated by the vaccination programs in British Columbia, across Canada, and in similar jurisdictions. Such jurisdictions have had a relatively small portion of the population naturally infected at the time of writing, and have begun vaccination with the very elderly during a time when social distancing measures have kept the reproduction number low while those over 80 years of age are vaccinated. Accordingly, we hold *R* at 1.05 from January 1, 2021 for 60 days. We retain the model structure of Bubar et al.^15^ which stops and restarts the simulation, moving individuals daily into the appropriate vaccination compartment. After 60 days we raise *R*, typically to 1.15, 1.3, 1.5 in the main text and with some higher examples in the supplement. This rise in transmission models either relaxation of distancing measures, reduced compliance to widespread distancing measures, or rising frequencies of higher-transmission COVID-19 variants of concern^22^. During the next 210 simulated days we proceed with the specified vaccination scenario. We model age-specific hesitancy, with some portion of each age group declining the vaccine.

We consider five vaccination scenarios. In all scenarios the 80+ age group is vaccinated first. In scenario A available vaccines are distributed to age groups in order of decreasing age. In scenario B, after the 80+ group is vaccinated, the vaccine is distributed to everyone else with no preference for age. In scenario C, D, and E, after the 80+ group, essential workers are then vaccinated without regard for age. In scenario C, the rest of the population is vaccinated in decreasing order of age. In scenario D, the rest of the population is vaccinated without regard for age. In scenario E, the 70–79 cohort is vaccinated next and then the rest of the population is vaccinated without regard to age.

For each vaccination scenario and choice of parameters, we measured multiple outcomes: number of infections, deaths, hospitalization, and cases of Long COVID. Whether any specific infection became a case of Long COVID was determined by an age-dependent probability which was computed using data from the Covid Symptoms Study App, (CSSA)^55^. The CSSA defines Long COVID as having symptoms longer than 28 days. Hospitalizations and deaths per detected case were estimated from data provided by the Public Health Agency of Canada^30^, Table 13-26-0003, for infections detected in the period between Sept. 15, 2020 and Jan. 15, 2021. At this time, the ascertainment fraction is believed to have been relatively high^23^. We use an ascertainment fraction of 0.75 to scale the estimated hospitalization and death rates per detected case to infections in the model, and compared the resulting hospitalizations and deaths (along with incidence) to data from British Columbia, Canada in fall 2020 (see Figure S1).

A disproportionate number of deaths have been among those living in long term care (LTC) settings, accounting for 2/3 of British Columbia’s deaths and 80% of Canada’s death in the earlier part of the pandemic^40, 59^ despite the fact that well under 10% of Canada’s seniors aged 65 and over are in long term care^4^. Long term care settings have been prioritized for vaccination and there are already indications of benefits. Accordingly, we reduced the infection fatality rate to reflect the fact that LTC settings are very unlikely to see death rates as high as they experienced in the pandemic to date; we reduced the rate by 1/3, consistent with the above numbers and the PHAC data on cases by age^47^.

Finally, we considered some economic measures of the cost of the pandemic from a health system payer perspective: Health utility losses measured in QALYs lost and NMB. Estimating health utility in terms of QALYs allows us to quantify and compare loss of quality and duration of life due to illness, disability, and death^60^. QALYs are estimated such that the maximum value of 1 indicates a year in perfect health, whereas a value of 0 indicates no health (death), and the measure is unbounded such that negative values capture health states worse than death. Utility decrements due to acute COVID infection, hospitalizations, chronic outcomes, and death are estimated as the difference between expected health, and health utility due to COVID. Our calculations built on the work of Briggs et al^12^, with additional material drawn from Kirwin et al^34^.

For each death due to COVID at a given age QALYs lost are the number of expected years of life left for someone of that age but with two adjustments. Firstly, since those who die of COVID are more likely to have preexisting conditions that shorten one’s life, we included a standardized mortality ratio^12^ (SMR) of 2. Secondly, we discounted future years of life lost by 1.5% per annum, following the guidelines of CADTH^36^. We did not assume a reduction in baseline quality of life for individuals who died from COVID.

For the loss of QALYs due to acute infection we used figures from Kirwin et al^34^. Each infection was given an age-dependent loss of quality adjusted days (Table 1). Hospitalizations incurred a utility decrement of 0.58, which was adjusted for mean duration of hospital stay by age group, to obtain the quality adjusted days lost for a hospitalization. For those discharged from the hospital, an additional utility decrement of 0.1 QALY was applied for the one-year period following discharge, as individuals do no immediately return to their pre-COVID health state following discharge^34^.

**Table 1:** Death, hospitalization, and Long COVID rates by age. Death and hospitalization rates are based on publicly available data from the Public Health Agency of Canada (PHAC; StatCan Table 13-26-0003)^30^ and Public Health Ontario^2^. Death rates for ages 0–49 were taken from the Ontario data because the PHAC values were zero. The death rate among those 80+ was adjusted to account for vaccination that has already taken place in long-term care. Rates of Long COVID were estimated from data in Sudre et al^55^, where Long COVID is defined here to be symptoms for more than 28 days. The proportion of essential workers by age was taken from the COVID Speak survey^3^.

| Age | Hosp rate | Death rate | Long COVID rate | Vaccine hesitancy | Prop. essential |
| --- | --- | --- | --- | --- | --- |
| 0–9 | 0.0054 | 2.00E-05 | 0.04 | NA | 0.0 |
| 10–19 | 0.0054 | 7.00E-05 | 0.04 | NA | 0.0 |
| 20–29 | 0.0110 | 3.1E-04 | 0.04 | 0.3 | 0.17 |
| 30–39 | 0.0203 | 8.4E-4 | 0.08 | 0.2 | 0.20 |
| 40–49 | 0.0283 | 0.0054 | 0.15 | 0.2 | 0.17 |
| 50–59 | 0.0557 | 0.0021 | 0.25 | 0.2 | 0.15 |
| 60–69 | 0.1280 | 0.0132 | 0.25 | 0.15 | 0.16 |
| 70–79 | 0.2750 | 0.0711 | 0.25 | 0.15 | 0.10 |
| 80+ | 0.3098 | 0.1468 | 0.25 | 0.15 | 0.0 |

**Table 2:** Model Parameters

| Parameter | Value | Source |
| --- | --- | --- |
| max duration of chronic outcome | 25 years |  |
| fraction of hospitalization leading to chronic outcome | 0.2 |  |
| utility decrement of chronic outcome | 0.16 | see SM |
| cost of hospitalization | \$1,391.88 | <sup>34</sup> |
| cost of year of chronic outcome | \$10,019.88 | see SM |
| dollar value per QALY | \$30,000 | <sup>36</sup> |

The QALY lost for chronic outcomes of COVID infection is the factor with the greatest uncertainty, since we have not been able to observe COVID survivors for multiple years. We started by estimating utility decrements for chronic outcomes of COVID (including those that fall under the rubric of Long COVID such as ME/CFS, and other complications). We identified all chronic conditions which have been observed at a higher rate among COVID hospitalization cases versus controls and collected utility decrements and annual incremental health system costs associated with each of these conditions. We weighted the collected values by the prevalence of each condition in the general population (pre-pandemic), and adjusted each estimate for duration of symptoms, survival, and discount rate. (For details see Supplemental Materials.) We estimated an annual utility decrement of 0.16. To approximate the (highly uncertain) future burden of chronic outcomes due to Long COVID and COVID complications, we assume that COVID infections that are severe enough to require hospitalization have a 20% chance of leading to a chronic outcome (whether it be ME/CFS or another) that has this utility decrement for whichever comes sooner, 25 years or the natural end of their life. This probably overestimates QALYs lost in some ways (maybe chronic outcomes improve after a few years, and/or perhaps they have more modest utility decrements) and an un-derestimate in others (one does not have to be hospitalized to get Long COVID). We then adjusted for currency and inflation, and discounted future years of life lost by 1.5% per annum as before. We intend this term to include all chronic outcomes of COVID-19 infections (both Long COVID and COVID complications) which likely include many conditions of varying frequency, duration, and severity.

We then determined a NMB (loss) for the pandemic, first by converting QALYs lost to a monetary equivalent value by multiplying by $30,000 Canadian dollar^36^ and then adding estimates of costs due to hospitalization and chronic outcomes. Using estimates of the direct costs of hospitalisation from Kirwin et al^34^ gave $1391.88 per day of hospitalization. The cost of chronic outcomes was again estimated by weighting the cost per year of the list of chronic conditions by their pre-pandemic prevalence, leading to a value of $10,019.88 per individual per individual annually.

Following the data from studies on the Pfizer and Moderna vaccines on the impact of vaccination on infection and on viral load, and data from the clinical trials on the impact on severe disease^11, 32, 41–43^, we assume a 90% effectiveness in preventing illness and death, and then vary the effectiveness in preventing infection from 0.6 to 0.9.

Figure S1 shows simulated incident cases, hospitalization and deaths compared to data from British Columbia (BC), Canada, during a period of slow but sustained growth at *R* = 1.2 in fall 2020. This illustrates that the model produces realistic age-structured simulations for BC.

## 3 Results

We find that vaccinating essential workers earlier gives large reductions in infections, hospitalizations, deaths, and instances of Long COVID (cases with symptoms lasting longer than 28 days), across a range of scenarios for transmission and vaccine efficacy. Except for deaths, results were similarly good with a strategy that vaccinates younger people sooner without targeting essential workers. However, with scenarios vaccinating the elderly later, they have very slightly higher rates of adverse outcomes, as expected, depending on how they are prioritized.

Figure 1 illustrates the high impact of including younger age groups sooner in the program than in an primarily oldest-first strategy (A), either through expanding to all age groups after those over 80 have been vaccinated (strategy B) or through vaccinating essential workers of any age group after those over 80 and then continuing from oldest to youngest (C), or expanding to all ages after essential workers (D). Vaccinating essential (high-contact) workers early has a strong effect, and can be done within the context of a broader oldest-first vaccination scheme. This does not require additional doses of vaccine. The shading in Figure 1 indicates which age groups are affected in four example simulations in the model, with four distinct vaccination strategies. We find that Long COVID impacts primarily those under 60, unlike hospitalization and death, which disproportionately affect the elderly.

**Figure 1:**
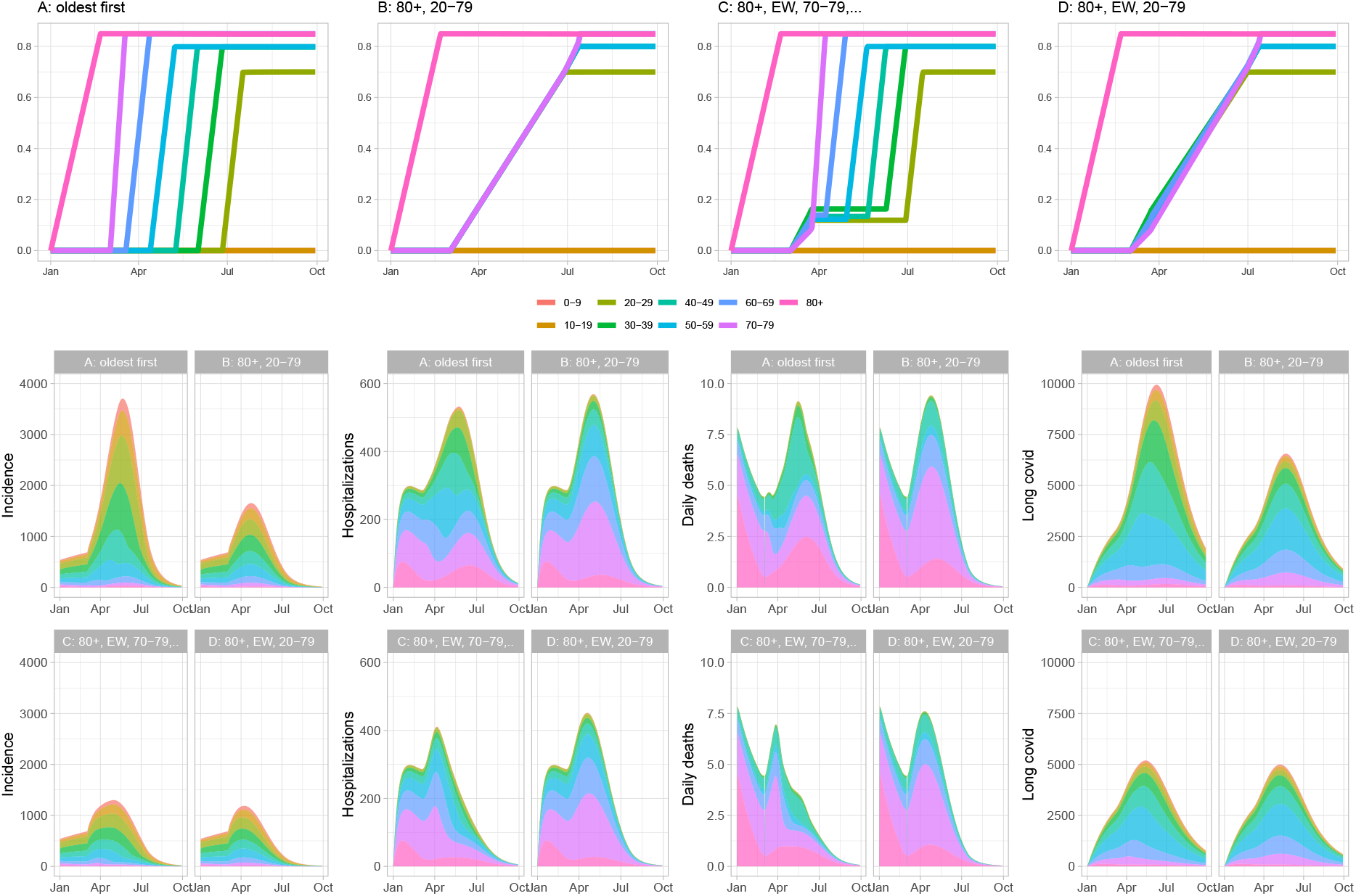
Vaccinating younger adults sooner, and vaccinating essential workers sooner, reduces cases, deaths, hospitalizations and Long COVID (COVID symptoms lasting longer than 28 days). In our model the rollout consists of an initial phase during which those over 80 are vaccinated and *R* = 1.05, followed by a second stage where the transmission rate is higher (*R* = 1.3 here). In these simulations vaccine efficiency is 90% for preventing disease and 75% at preventing infection and therefore subsequent transmission. Top row: illustration of the vaccination programs by age group over time. Next two rows: simulated cases, hospitalization, deaths and Long COVID prevalence over time.

Figure 2 shows the simulated total cases, deaths, hospitalizations and Long COVID cases for a range of transmission-blocking efficacy and for two values of the underlying *R*. At the time of writing, many jurisdictions are vaccinating on a primarily oldest-to-youngest vaccination program^5, 9, 29^, although some health care workers and staff in long term care homes and some other vulnerable communities are included in the early stages. In our model, this approach leads to con-siderably more cases and more Long COVID, across a range of values of the vaccine’s efficacy against transmission and across a range of *R* values. When *R* is kept very low (1.15) for example through continued strong social distancing, all of the simulated strategies do well at reducing deaths. When *R* rises to 1.3, strategies placing essential workers after 80+, either continuing with an age-based rollout or opening to all adults aged 20-69 after those 70+, have an advantage in reducing deaths in addition to strong advantages for infections, hospitalizations and Long COVID. Other differences between the age ordering are very small compared to the difference between “oldest first” strategies and *any* alternative that prioritizes essential workers or even all younger adults earlier in the program.

**Figure 2:**
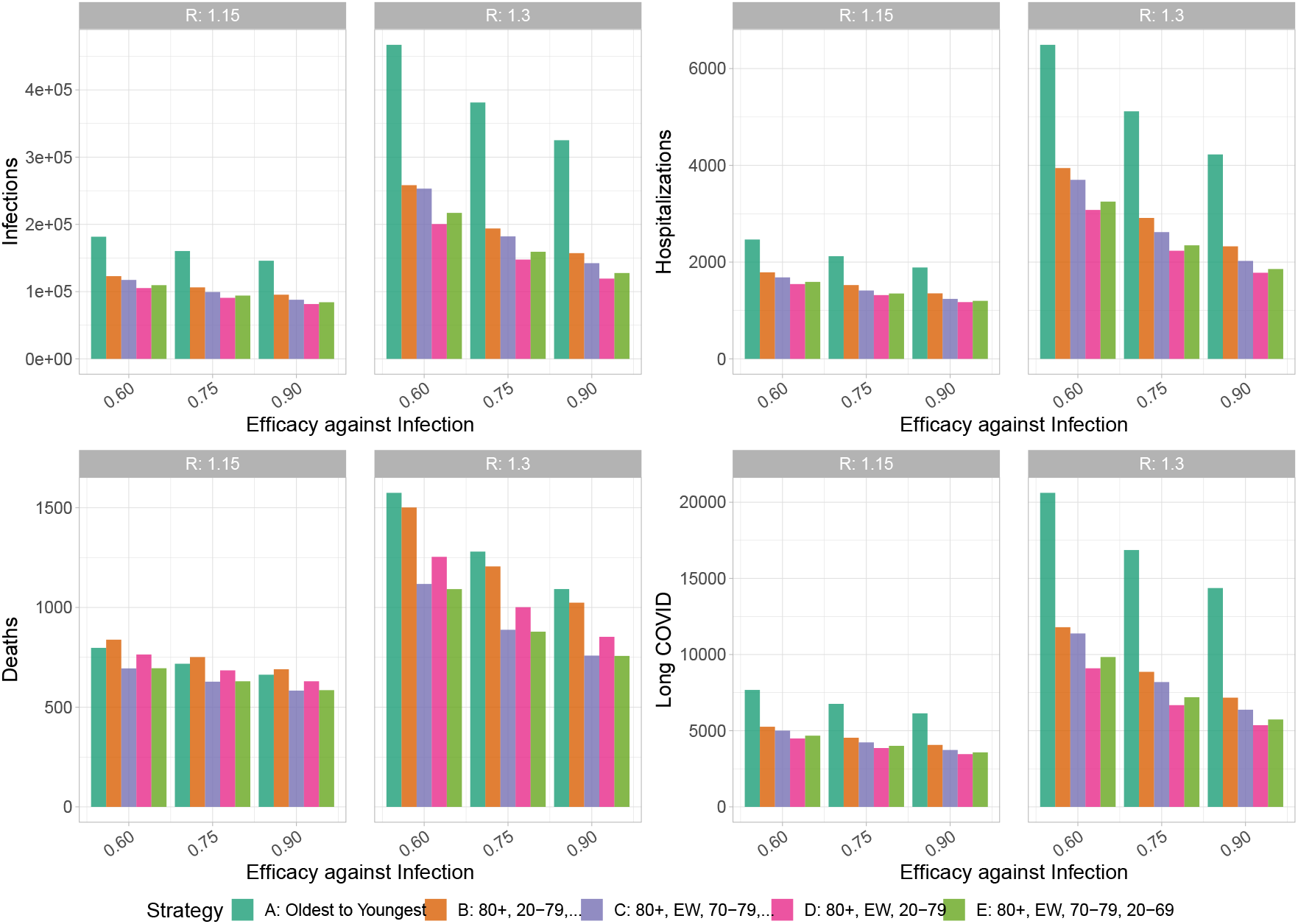
Vaccinating essential workers earlier in the program has benefits for cases, hospitalizations, death and Long COVID (COVID symptoms for longer than 28 days). This does not depend sensitively on the efficacy of the vaccine against transmission, nor on the underlying transmission rate. Similar results are attained by vaccinating over all age groups (strategy B), with the exception that this gives less reduction in deaths.

We explored the sensitivity of our vaccine strategy comparisons to the portion of workplace contacts taking place among essential workers (Figure S6), the efficacy against transmission (Figure S7), the portion of workers considered “essential” (Figure S8) and to the contact matrix (Figure S9). We explore a wider set of strategies (Figure S3). Consistently, vaccinating essential workers earlier has considerable benefits compared to oldest-first strategies while the relative merits of different age-based rollouts depend on assumptions. For example, when the portion of workplace contacts among nonessential workers is higher, strategy C (80+, EW, 70-79,..) which is predomi-nantly oldest first loses some of its benefit because essential workers have less of the overall contact (Figure S6). While the efficacy against transmission has a high impact on the outcomes, it does not much impact the relative performance of the strategies unless *R* is high (Figure S7). Finally, we determine the optimal strategy simply from point of view of minimizing deaths; despite the fact that the mortality risk motivates oldest-first vaccination, this strategy is only the best for deaths as an outcome when efficacy against transmission is extremely low (0.1-0.2) and when *R* is high (Figures S10 and S11). See the Supplemental Materials for further details.

Figure 3 shows health utility losses in QALYs and their source (cases, chronic impacts, deaths and hospitalizations). Here chronic impacts includes both chronic Long COVID symptoms (similar to ME/CFS) and other long-term complications. Deaths are naturally a large contribution to health utility loss; the next biggest contribution by far are the chronic impacts where we find that vaccinating essential workers sooner has profound benefits For example, if *R* rises to 1.3 and the vaccine is 75% effective in preventing transmission, the combined reduction in deaths, chronic impacts, cases and hospitalizations when essential workers are vaccinated after those aged 80+ means that over 11000 QALYs are gained (Figure 3, bottom middle panel). Of these, 3000 are due to chronic impacts. When efficacy against transmission is lower, because there are more infections, it remains very beneficial from a health utility point of view to vaccinate essential workers sooner. As with hospitalization, deaths and Long COVID, the differences between age rollouts beyond whether essential workers are included early is relatively small.

**Figure 3:**
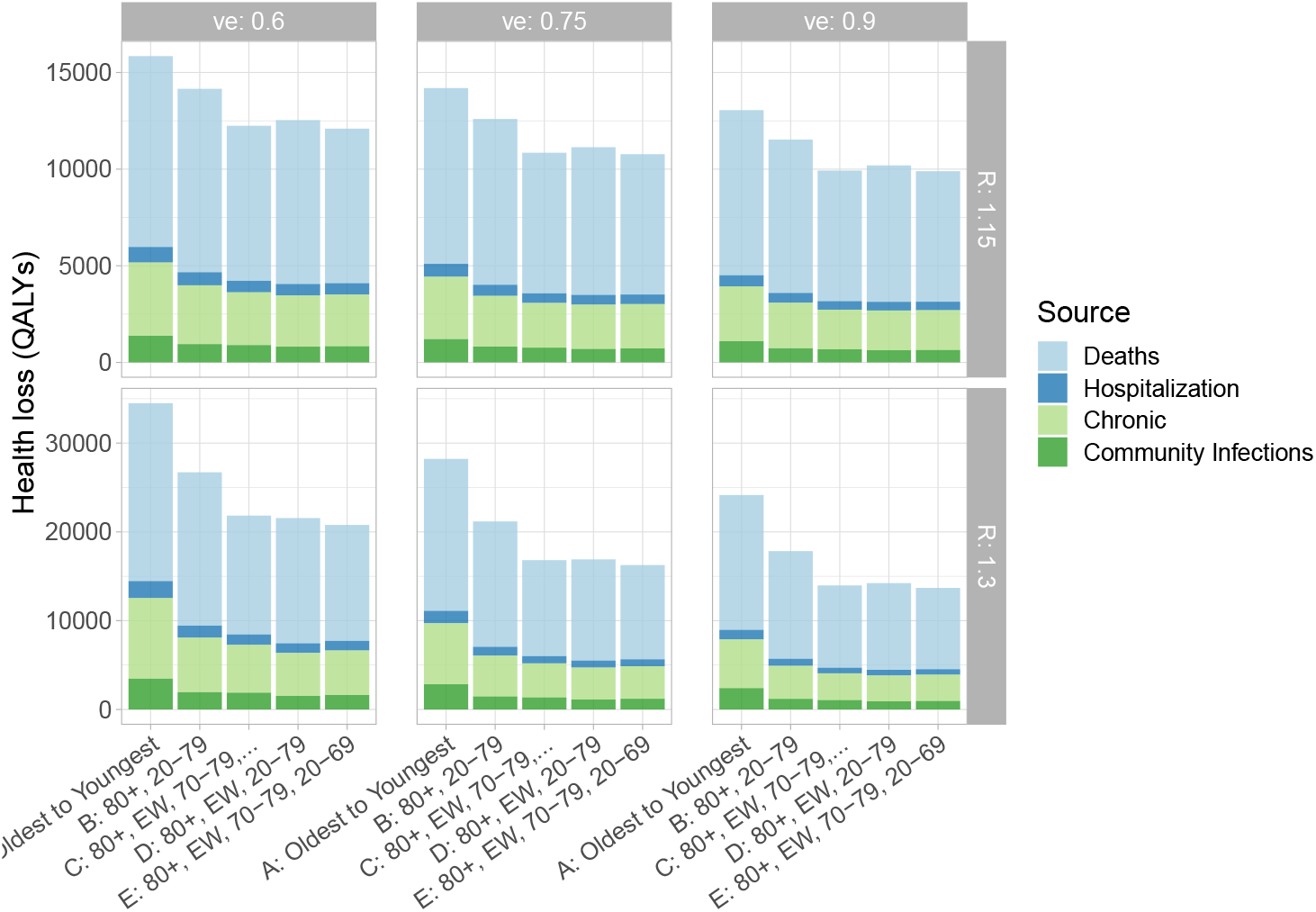
Health losses (QALYs). *R* and the efficacy against transmission vary and are labelled on the side (*R*) and the top (efficacy).

We find similar effects when we estimate the incremental costs of the pandemic including both direct costs of hospitalization and chronic outcomes and health utility decrements converted to monetary values (Figure 4). Vaccinating essential workers sooner reduces the overall NMB loss due to the pandemic by %50 to %65. This results in a potential savings of (for example) over $M400 (if *R* = 1.3); see Figure S2. In all scenarios, NMB lost was largest for the age-based immunization strategy (A). The largest improvement in NMB was achieved with strateges C, D and E, irrespective of vaccine efficacy. Figure 4 highlights the potential for a substantial impact of chronic consequences of infection, both related to health utility losses, and future health system cost.

**Figure 4:**
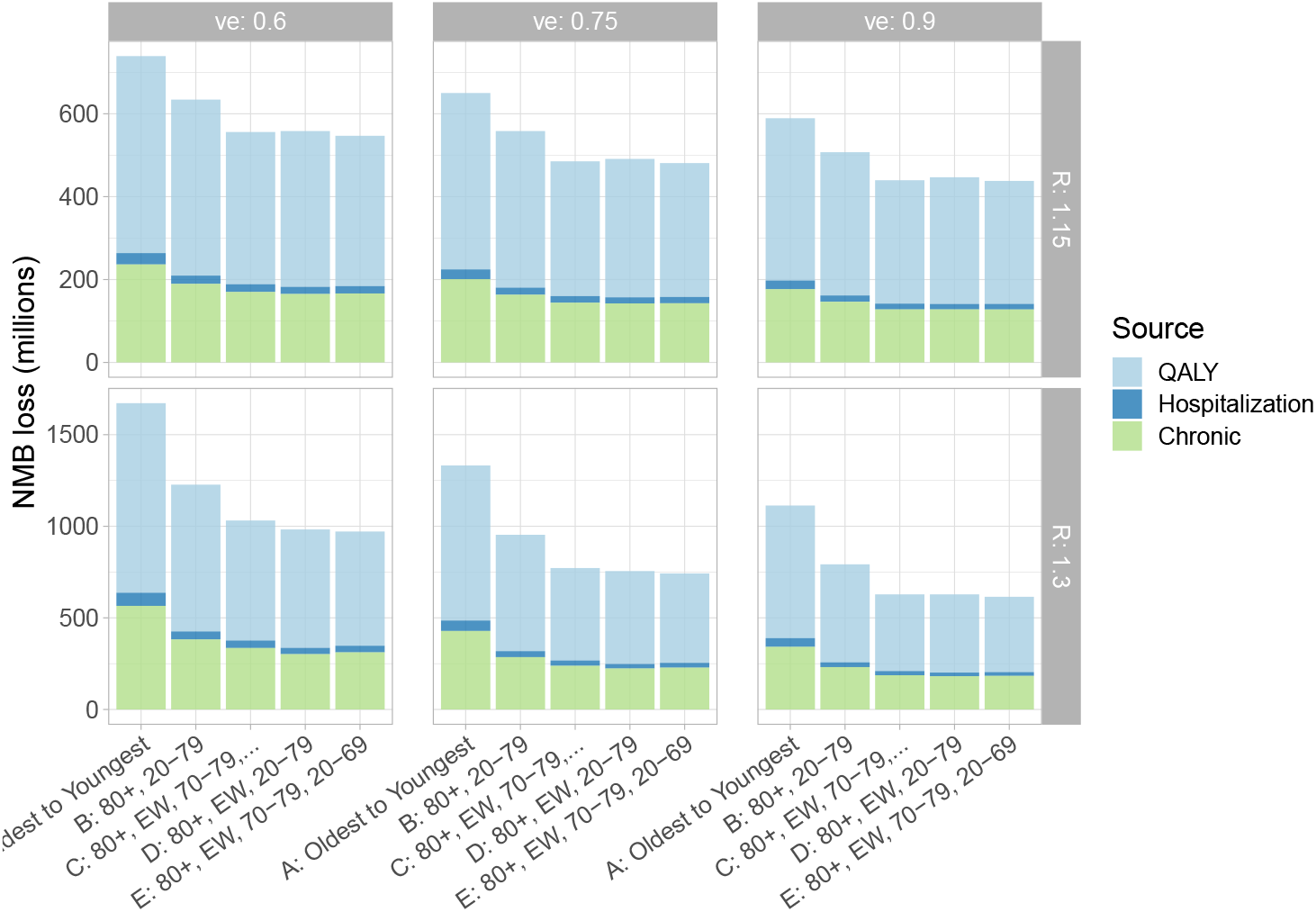
Net Monetary Benefit (loss), due to the cost of treating chronic outcomes, hospitalizations during the pandemic, and health utilty converted from QALYs lost shown in Figure 3. Transmission-blocking efficacy *𝓋e* and *R* vary as labelled over the panels.

Finally, we explored immunity as transmission began to decline (“herd immunity”). Figure 5 shows the fractions of the population who were infected (top panels), or who were protected by either natural infection or vaccination (bottom panels), at the time when simulated infections began to decline, for three transmission-preventing efficacies and for a range of *R*. Consistently, the oldest to youngest strategy requires more immunity than strategies vaccinating essential workers or younger people sooner. This is because individuals who have a comparatively low likelihood of exposure and transmission are vaccinated, but their protection contributes less to the population’s collective protection than vaccination of those who are at risk of exposure and transmission. Accordingly, vaccinating 80+, essential workers and then all ages requires the fewest infections and the least immunity in order for cases to decline; in this strategy those likely to transmit are protected efficiently. The immunity required for infections to decline is more sensitive to the vaccination strategy when *R* is relatively low; after *R* = 2, the required protection for all strategies is just under 1*/*2. However, vaccinating essential workers early allows far more of that fraction to be protected by vaccination, whereas in the oldest-first strategy more infection is required to achieve declining infections.

**Figure 5:**
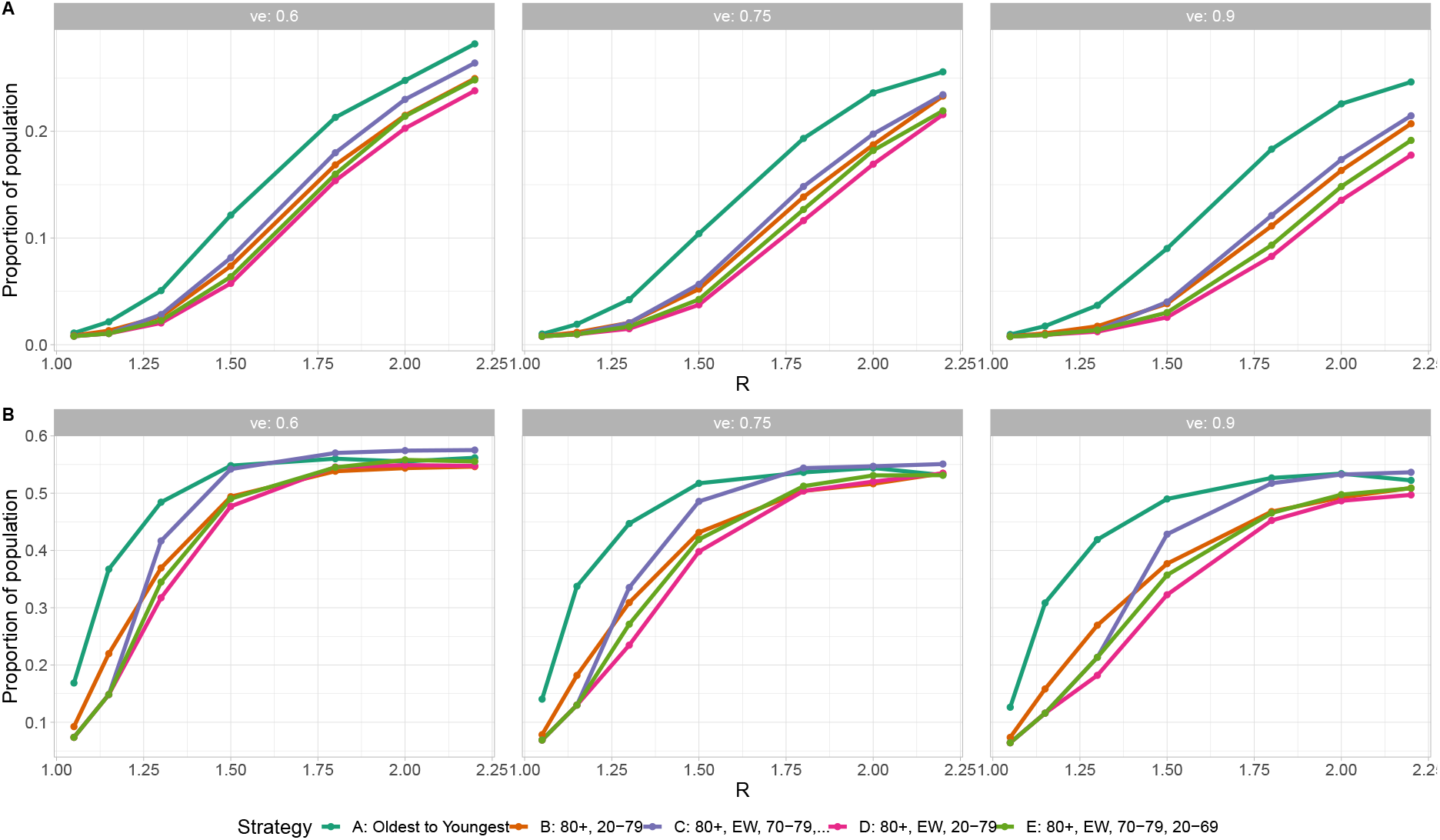
Immunity and vaccination at the time when infections begin to decline, as a function of *R*. (A) Proportion ever infected by turn around time. (B) Proportion ever infected or vaccinated by turn around time. The efficacy of vaccination against infection (*𝓋*_*e*_) is varied from 0.6 (left) to 0.9 (right).

## 4 Discussion

Strategies that vaccinate essential workers early lead to substantial reductions in the number of infections, hospitalizations, deaths, and cases of Long COVID relative to a strategy of oldest first. This finding is robust to details of how the vaccine is rolled out to younger people, the exact fraction of essential workers vaccinated, the effectiveness of the vaccine at preventing transmission, and the underlying transmission (described by *R*). The reason is that essential workers have many more contacts than older adults and vaccinating them is the most effective way of reducing overall prevalence. For the range of parameters we considered, the benefit of vaccinating essential workers and thereby reducing prevalence by preventing transmission outweighs the benefit of vaccinating older adults first. Reduction in deaths were often modest, but reduction in other measures were substantial, since the total number of infections was reduced. Benefits were similar even if instead of essential workers, young people were vaccinated along with older people irrespective of their work status, since young people tend to have a higher number of contacts on average.

By essential workers we mean people who have to have high contact as part of their job. This is distinct from “essential services” and beyond health care workers could include teachers, taxi drivers, retail workers, food production workers, law enforcement and public safety, first responders, social workers, agriculture, transportation and many more^26^. Our results hold for any group of similarly high-contact individuals; we focused on essential workers since these individuals cannot effectively isolate under any social distancing regime, and there is added moral importance to vaccinating them.

There is an important equity consideration, as the COVID-19 pandemic has disproportionately impacted essential workers. Across neighbourhoods in Toronto, for example, per-capita COVID cases and deaths were 2.5 to 3 times higher in neighbourhoods with high (vs low) concentrations of essential workers^48^. Essential workers often have lower incomes, and may be hired as contractors; they may not have paid sick leave, and may have limited ability to negotiate safe working conditions^48^. While many others are able to work safely at home, these individuals cannot. Our findings suggest that prioritizing them for vaccination not only would help to reduce this substantial disparity, it does not even come at a cost of increased adverse outcomes in others; rather, it is better for everyone.

Our reason to consider low values of *R* exclusively is because that is the condition most jurisdictions in Canada, the US, and Europe operate at, i.e. with *R* relatively close to 1. Larger values lead to the rapid overwhelming of the healthcare system, and governments inevitably introduce lockdown or stricter social distancing measures in response. For example, across Canada, lockdown measures were introduced when incidence reached approximately 20-50 cases per 100K population per day (or 1000 cases per day in BC). *R* = 2 in the model results in incidence 40–60 times that. Even at a reduced effective hospitalization rate (with older groups vaccinated), the health care system would be overwhelmed if these incidence levels were reached. Accordingly, *R* is not likely to remain high enough for oldest-first strategies to be best; distancing and other measures have consistently been put in place to prevent this.

We modelled a rise in transmission 60 days into the simulation, in part because of increasing evidence that variants of concern (VOC) are rising in frequency in Canada^14^ and elsewhere^58^. A number of distinct VOC have emerged, showing signs of increased transmissibility^7^, and there have already been introductions and community exposures across Canada. As their frequency rises, VOC are likely to drive higher transmission rates, particularly in the current context where many areas are considering relaxing restrictions following declining cases in recent weeks. If VOC transmission is contained, the relaxation of measures itself is likely to result in higher transmission in coming weeks.

We explore the portion of the population who are immune at the time that transmission begins to decline and find that the levels of protection at this “herd immunity” time are comparable to the theoretical requirement (1 *−*1*/R*) in very simple models, despite the fact that the model has heterogeneous contacts by age and work patterns. This is in contrast to other modelling work which has suggested that heterogeneous contacts mean that far fewer individuals might need to be protected^8, 13^. We note that unlike these studies, our model was compared to age-stratified incidence, hospitalization and death data and matched well. The difference is driven by the fact that our model has less severe contact heterogeneity than these models, and so has sufficient intergroup mixing that relatively high levels of immunity are required for herd protection.

The NMB results provide a synthesized measure combining several outcomes: infections, hospitalizations, deaths, and chronic outcomes of infection. This addresses challenges related to identifying appropriate vaccination program objectives: for example, it is not necessary to choose between optimizing reductions in deaths compared to reductions in hospitalizations. We note that we did not account for the costs of maintaining the low transmission levels throughout the duration of the simulation, and accordingly, lower *R* results in lower lost NMB. Our NMB losses are intended to compare vaccination strategies within a social and policy framework determining the levels of distancing and COVID-19 transmission.

Even without considering Long COVID and COVID complications, vaccinating essential workers sooner has strong benefits in terms of reducing infections, hospitalizations, deaths, and in terms of net monetary benefit. However, taking chronic outcomes into account makes the advantages of our proposed vaccination schedules over oldest-first even more stark, showing that they potentially save hundreds of millions dollars of additional NMB. These long-term consequences of COVID infection could impact future health of 0.5% of the population under an oldest-first vaccination strategy, and far fewer (0.25%) if essential workers and/or younger adults are vaccinated earlier. Despite uncertainty in the likelihood and duration of long-term consequences of COVID infection, Long COVID and COVID complications need to be included in considerations of vaccine priority.

## Data Availability

All data is available on a GitHub repository.

https://github.com/nmulberry/essential-workers-vaccine/

## Supplemental Materials

### Supplementary Results

#### Validation

In Figure S1, we validate the model contact structure, and the age-based hospitalization and fatality rates against observed cases, hospitalizations, and deaths in British Columbia over a period from October 1 to December 1 2020.

#### Net Monetary Benefit relative to Oldest-to-Youngest

In Figure S2 we show the Net Monetary Benefit of each of the alternative vaccination strategies, relative to strategy A (vaccinating from oldest to youngest). The NMB of any of the strategies relative to no vaccinations at all is considerably larger^2^.

Figure S3 explores two additional strategies that were not considered in the main text. Both of these additional strategies prioritize essential workers. Strategy F is similar to strategy E, except we consider allocating vaccines to 70–79-year-olds before essential workers (followed by a general rollout). Strategy G is again similar to strategies C and E, where we prioritize essential workers, then target older adults sequentially, followed by all 20–59-year-olds. We see that all of strategies C through G (which prioritize essential workers) out-perform the purely age-based strategies (A and B) in terms of infections, hospitalizations and deaths. However, the strategies which also prioritize adults aged 70–79 (strategies E–G) outperform strategy D (which does not prioritize this age group) in terms of mortality. Across outcomes, we observe that strategies E–G perform very similarly.

### Efficacy of the vaccine against Long COVID

In our model, individuals with Long COVID do not continue to transmit COVID, and their numbers therefore do not affect the dynamics of the rest of the model. Here we illustrate the impact of assumptions about the vaccine’s efficacy against Long COVID.

The model in the main text used the same efficacy against Long COVID as for severe outcomes (0.9). Figure S4 illustrates that this efficacy has a minor outcome on the Long COVID prevalence. Although we explore an efficacy of 0.6 compared to the default of 0.9, so a 30% decrease in efficacy this has a minor impact (a less than 30% reduction in Long COVID), because most of the infections in these scenarios occur among those who were not vaccinated. Most of the Long COVID cases also, therefore, occur among the unvaccinated.

### Sensitivity Analysis

We explore the sensitivity of our main result—that strategies which prioritize essential workers outperform age-based only strategies—to various model parameters. We show that our conclusion on prioritizing essential workers is robust (except in a small, and perhaps unrealistic parts of the parameter space), but that distinguishing among such strategies is often sensitive to various model parameters.

First, we explore transmission rates that so far seem unrealistic for the Canadian setting but have been used in other models^1^. Figure S5 shows trajectories where *R* is 1.5 or 2, vaccination proceeds at 0.4% per day with an efficacy against transmission of 0.6 and efficacy against disease is at 0.9 as in the main text.

Next, we explore the sensitivity of our results to *α*, the fraction of workplace contact occurring among nonessential workers (taken the be zero in the main text). Figure S6 shows that for values of *R* between 1.15 and 1.5, our conclusion that prioritizing essential workers outperforms an age-based rollout is robust to changes in *α*. However, as *α* increases the net-benefit of targeting essential workers (strategies C–E) over younger adults (strategy B) decreases (note that when *α* = 1, all working age adults have the same level of contact, regardless of essential worker status). Therefore, the relative performance of strategies which target essential workers (strategies C–E) is sensitive to both *R* and *α*, although our conclusion that such strategies outperform general age-based rollouts is robust.

We similarly explore the sensitivity of our results to a broader range of *𝓋*_*e*_, the efficacy of vaccination against infection, in Figure S7. We find that strategies which target younger adults sooner (strategies B and D) perform poorly in terms of deaths when vaccine efficacy is very low (*𝓋*_*e*_ *<* 0.5) across the different *R* values. Strategies that target essential workers and also older adult age groups (strategies C and E) are robust even for low values of *𝓋*_*e*_.

We furthermore explore the sensitivity of our results to changes in the contact matrix structure. First, we consider changing the proportion of essential workers (Figure S8). We find that strategies C and D, which prioritize essential workers and also older adults, are robust to changes in this frequency. However, strategy D (which does not prioritize older adults) is sensitive to changes in this frequency. Next, we find that our results are not qualitatively altered by random perturbations in the contact matrix (Figure S9). However, these results highlight that we may not be able to use this model to determine the small differences among the benefits of strategies C–E (all of which target essential workers, but differ in the details of the rest of the rollout).

Finally, in Figures S10 and S11 we test the relative performance of strategy C (which targets essential workers) against the age-based only strategies (A and B) over a wide range of parameters. These figures show the optimal strategy in terms of minimizing mortality for the given set of parameters. We find that strategy A (oldest to youngest) is only optimal in a very small region of the parameter space. Strategy C, which targets essential workers, is optimal almost everywhere. We note that we do not distinguish here across strategies D–G, which also target essential workers. These figures show that our conclusion that age-based only rollouts are non-optimal is robust except in parts of the parameter space which we consider to be unrealistic.

**Figure S1:**
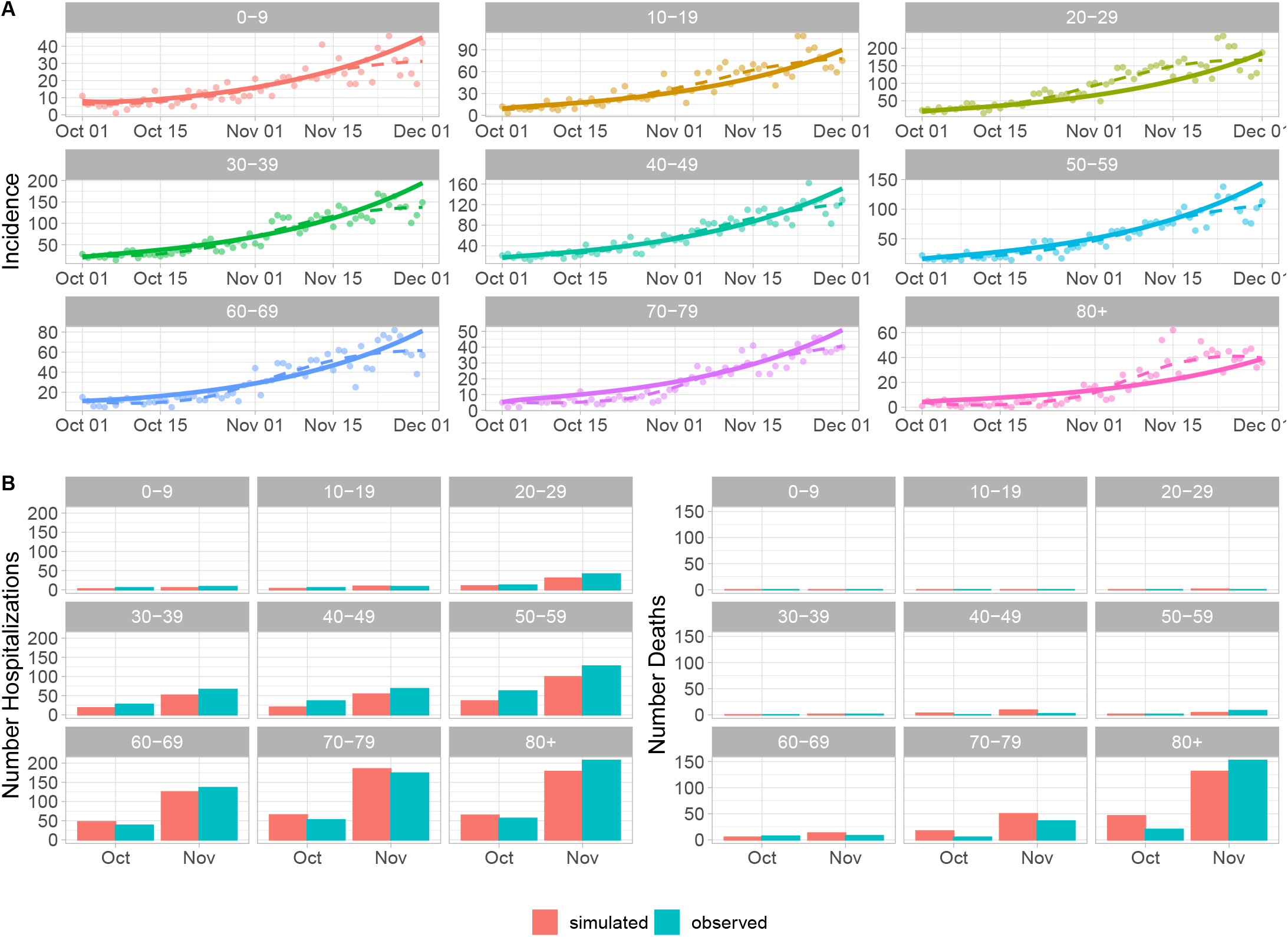
Validation with the age distribution of cases, hospitalizations and deaths. A) Dots represent actual reported case counts by age; lines are simulated values. B) Bars represent total hospitalizations and deaths by month. Time period Oct-Dec 2020. Constant *R* = 1.3. Initial conditions taken from case count data.

**Figure S2:**
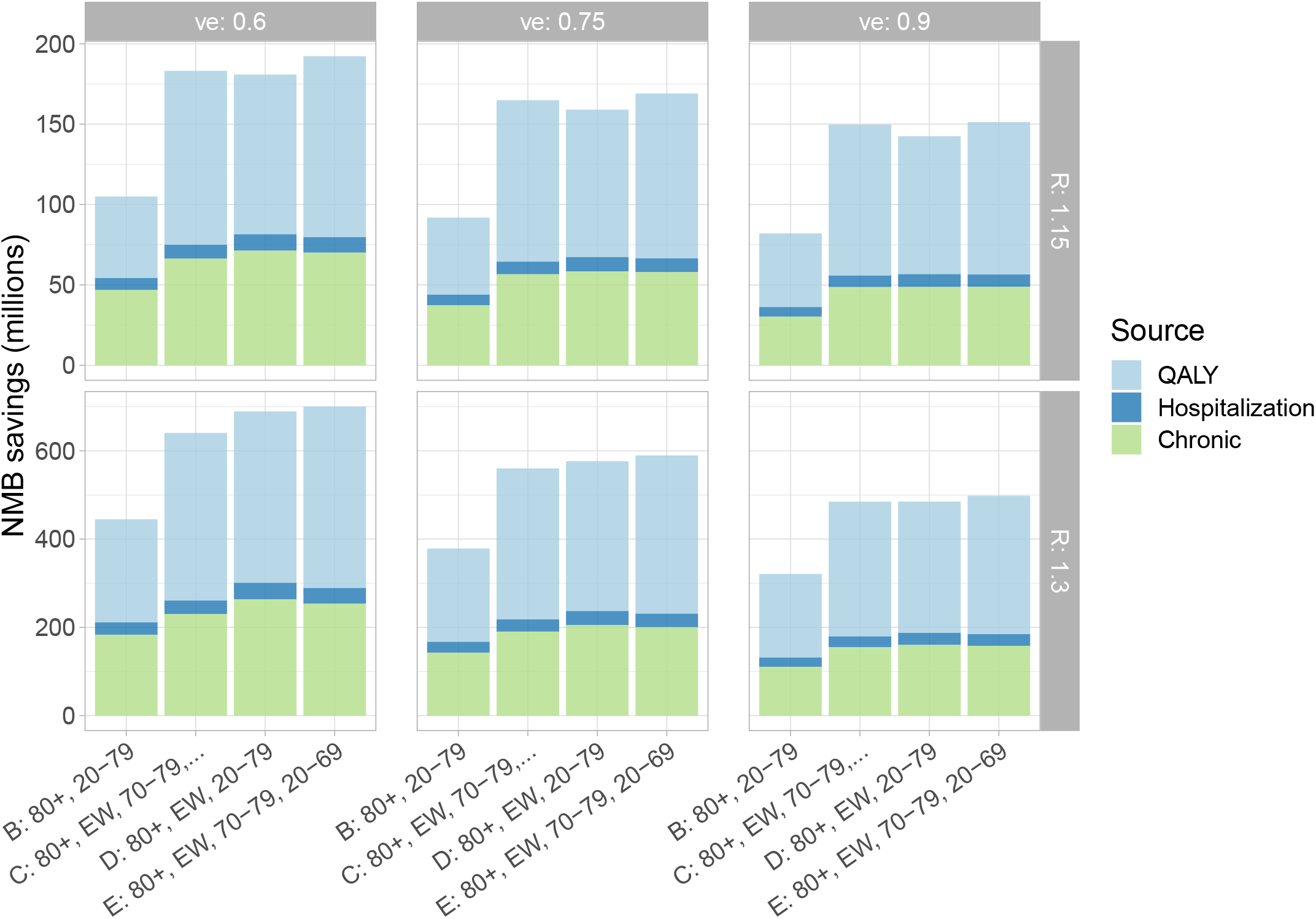
Savings for each strategy in millions, compared to Oldest to Youngest (strategy A). The parameter *𝓋*_*e*_ denotes the efficacy against transmission (top labels) and *R* is 1.15 (top) or 1.3 (bottom).

**Figure S3:**
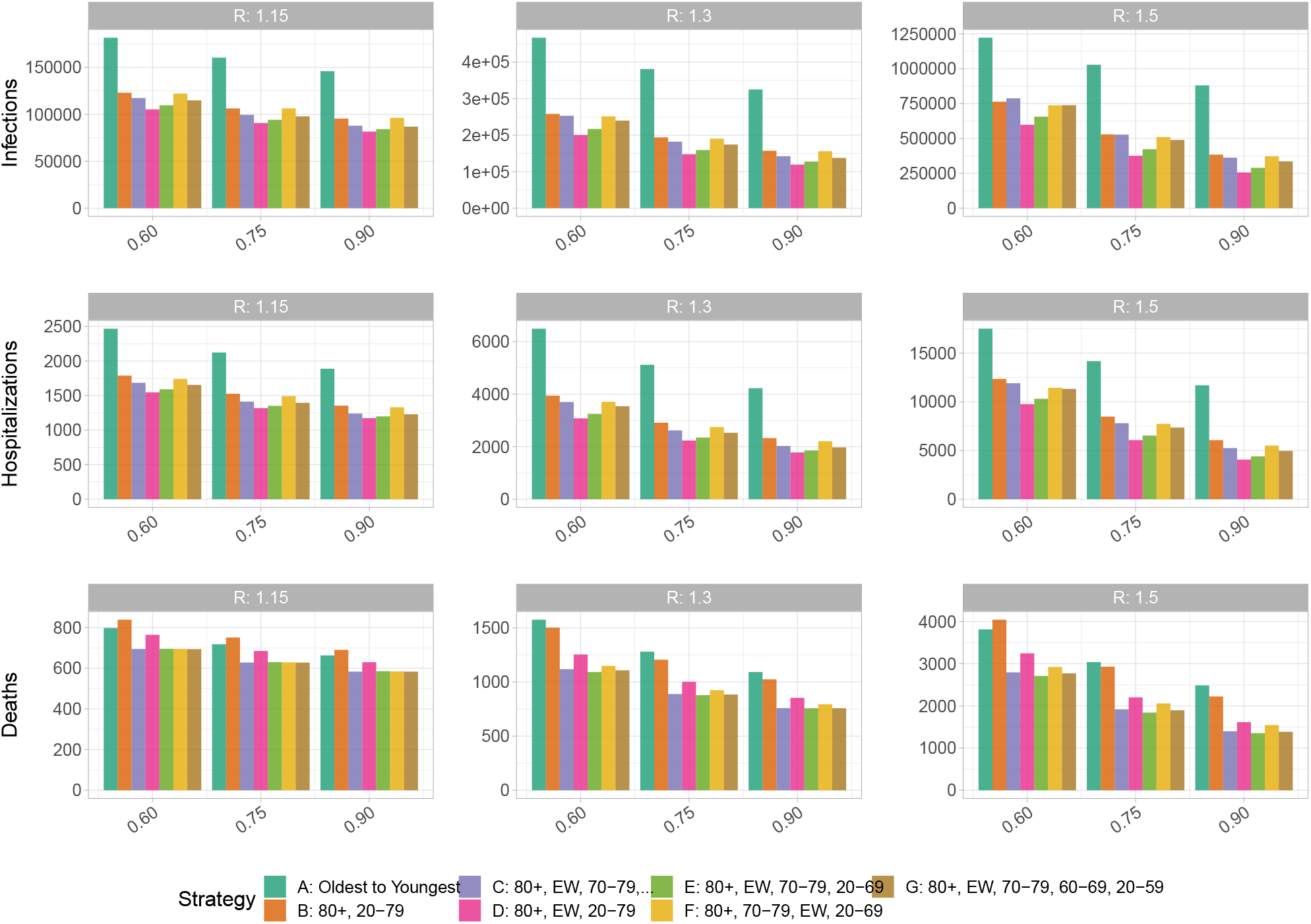
Alternative strategies. Fixed *α* = 0, *υ*_*p*_ = 0:9.

**Figure S4:**
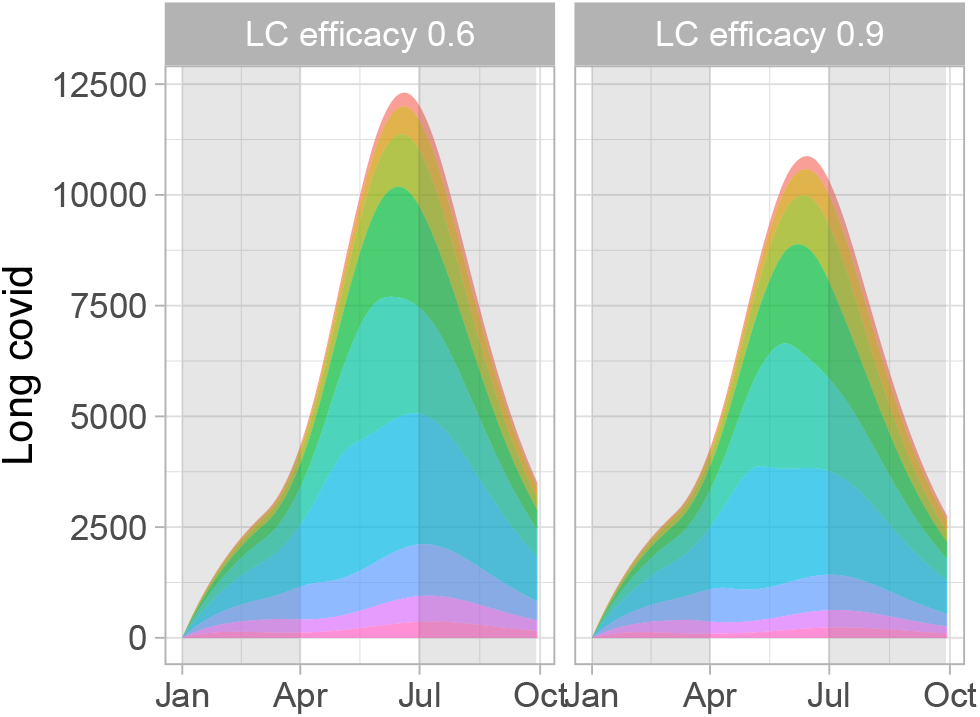
The Long COVID prevalence over time is not impacted much by a substantial change in the vaccine’s efficacy against Long COVID. In this demonstrative example, *R* is 1.05 during vaccination of those over 80, and then rises to 1.3 reflecting reopening or rising frequency of a higher-transmission variant. Colour indicates age as in Figure 1.

**Figure S5:**
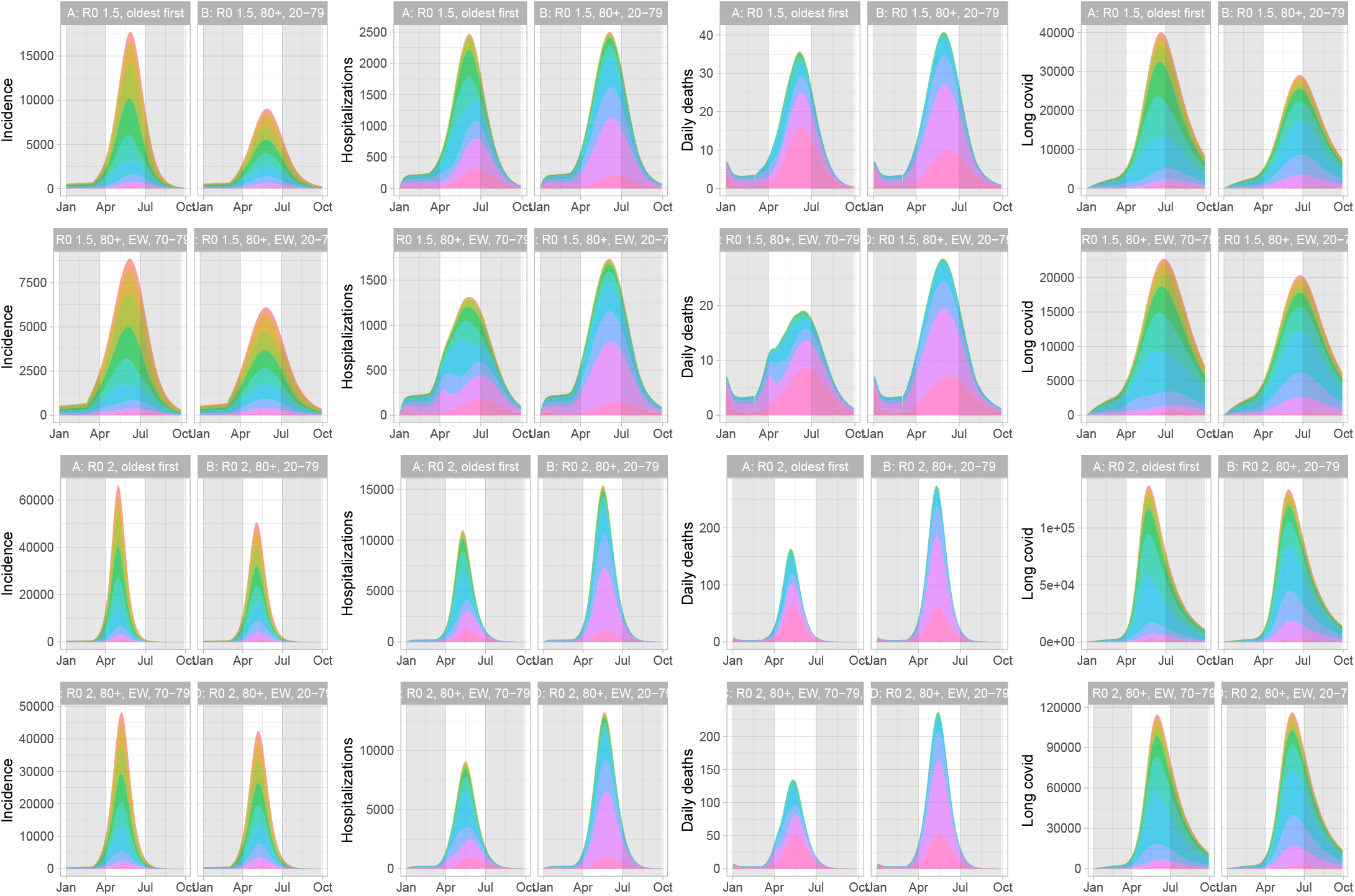
Trajectories for a higher *R* value (1.5, 2). Here, vaccinating younger people sooner still has a benefit for infections and Long COVID, but results in higher deaths and hospitalizations. However, vaccinating essential workers in the context of an age-based rollout reduces all negative outcomes substantially when *R* = 1.5, and has limited impact when *R* = 2.

**Figure S6:**
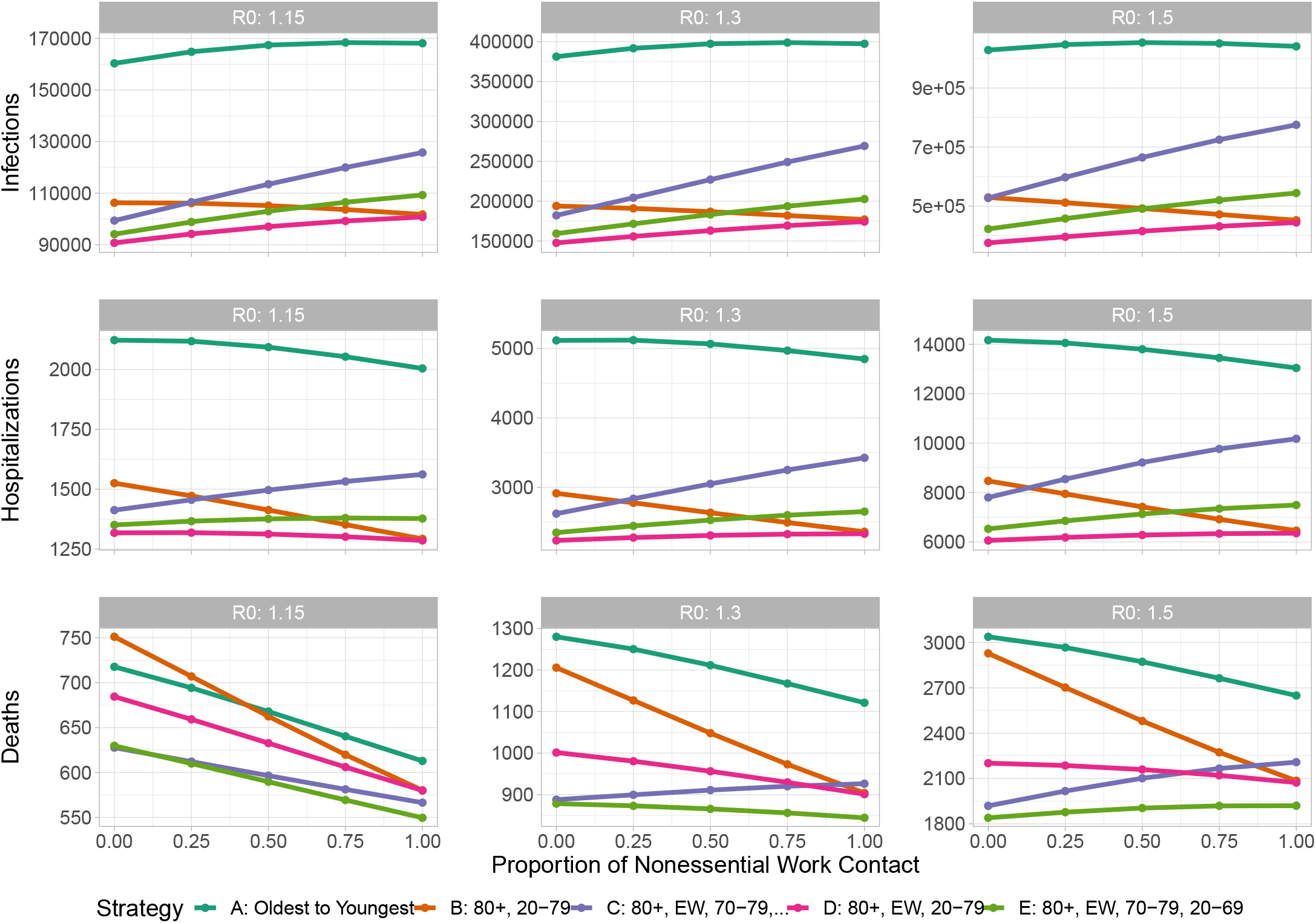
Sensitivity with respect to *α* (the proportion of workplace contacts among nonessential workers). We vary *R* from 1.15 (left) to 1.5 (right). We fix *𝓋*_*e*_ = 0.75, *𝓋*_*p*_ = 0.9. The oldest-to-youngest strategy consistently performs worse than the others. The relative benefits of the other strategies are typically small and depend somewhat on *α*, though strategies including essential workers early (pink, green) consistently do well.

**Figure S7:**
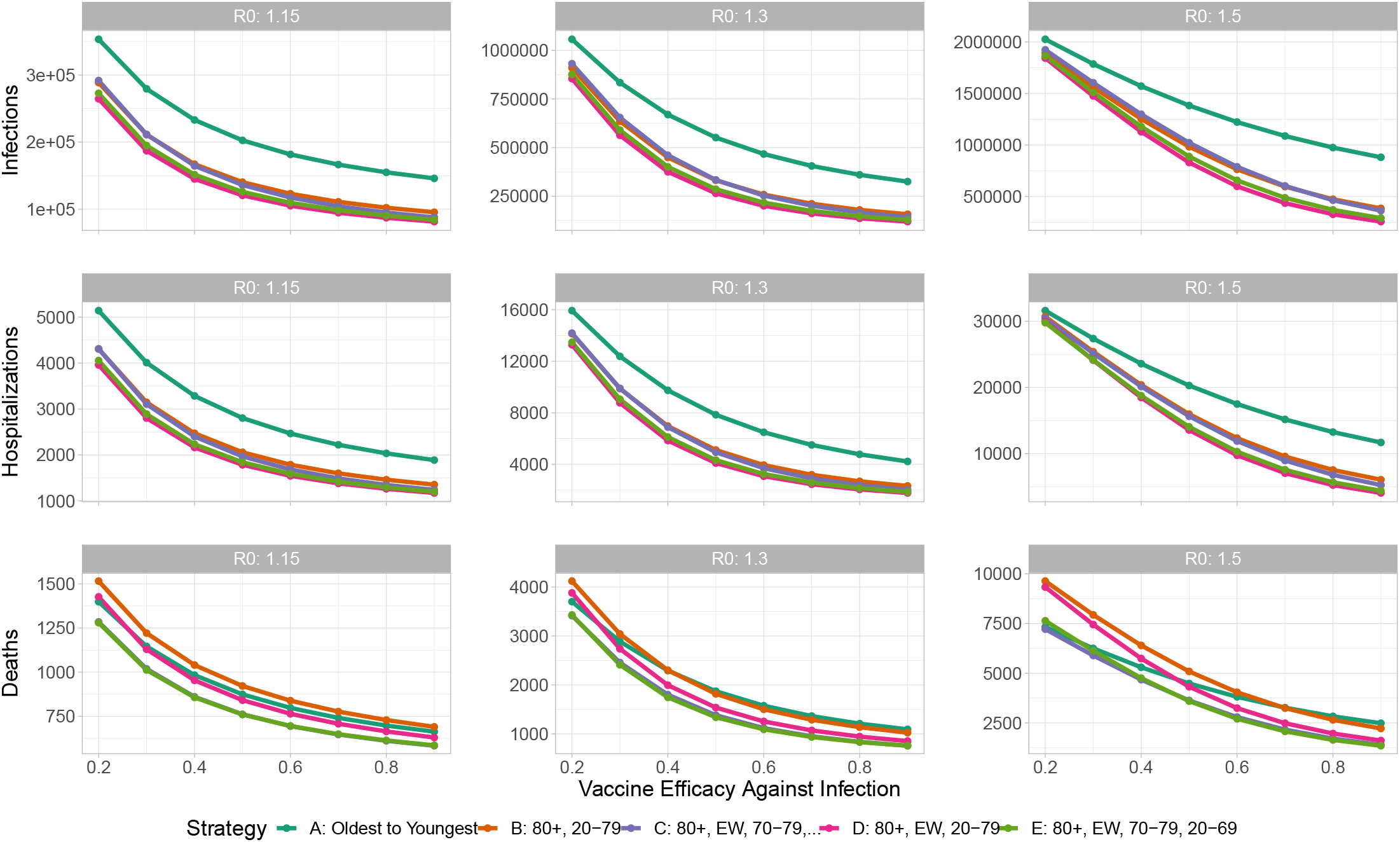
Sensitivity with respect to *𝓋*_*e*_ (the efficacy of vaccination against infection). We vary *R* from 1.15 (left) to 1.5 (right). We fix *α* = 0, and *𝓋*_*p*_ = 0.9.

**Figure S8:**
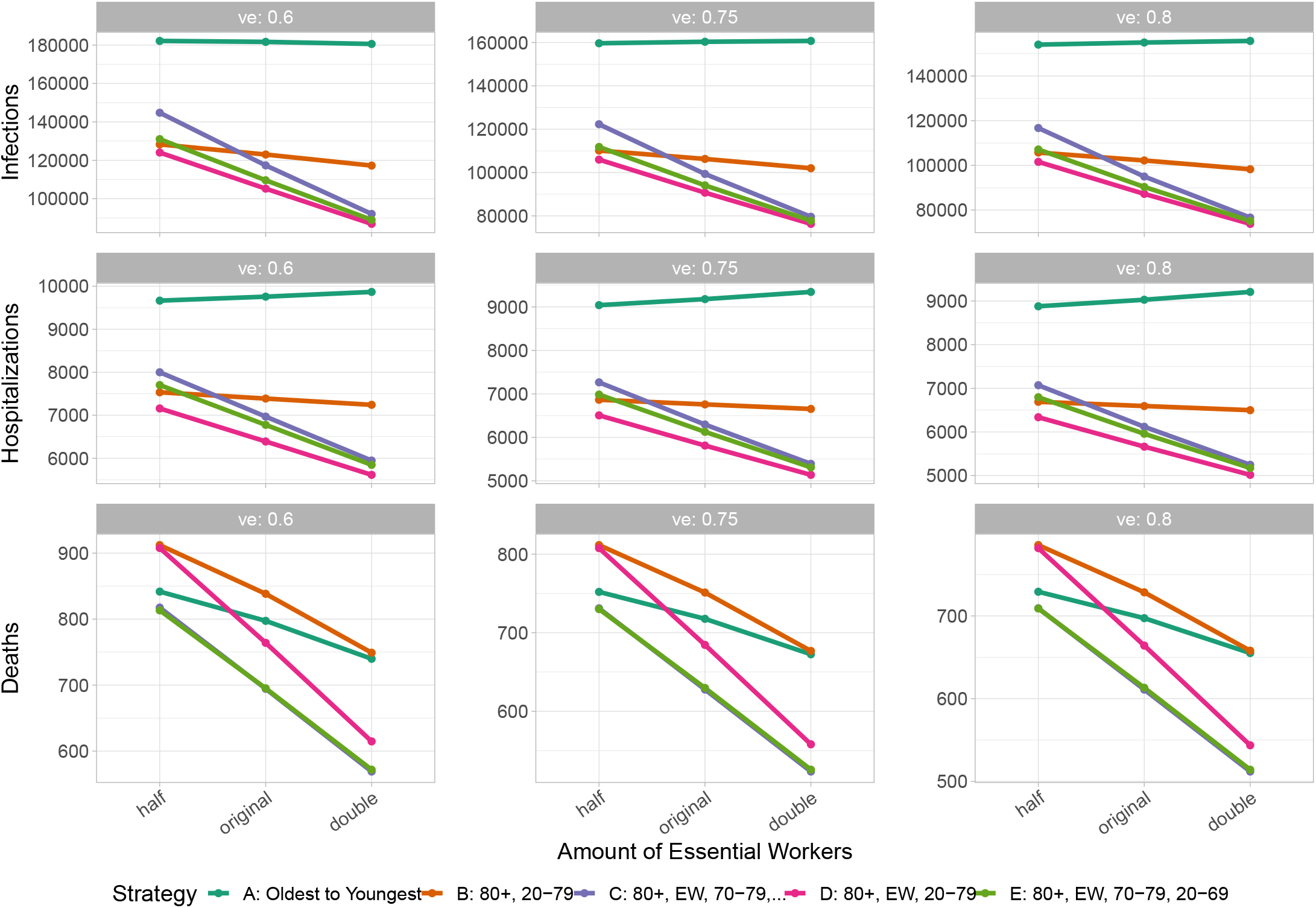
Sensitivity with respect to the proportion of essential workers. We vary *𝓋*_*e*_ from 0.6 (left) to 0.8 (right). We fix *α* = 0, *𝓋*_*p*_ = 0.9, *R* = 1.15. Similar results for other values of *R*. Note that strategies C and D often overlap.

**Figure S9:**
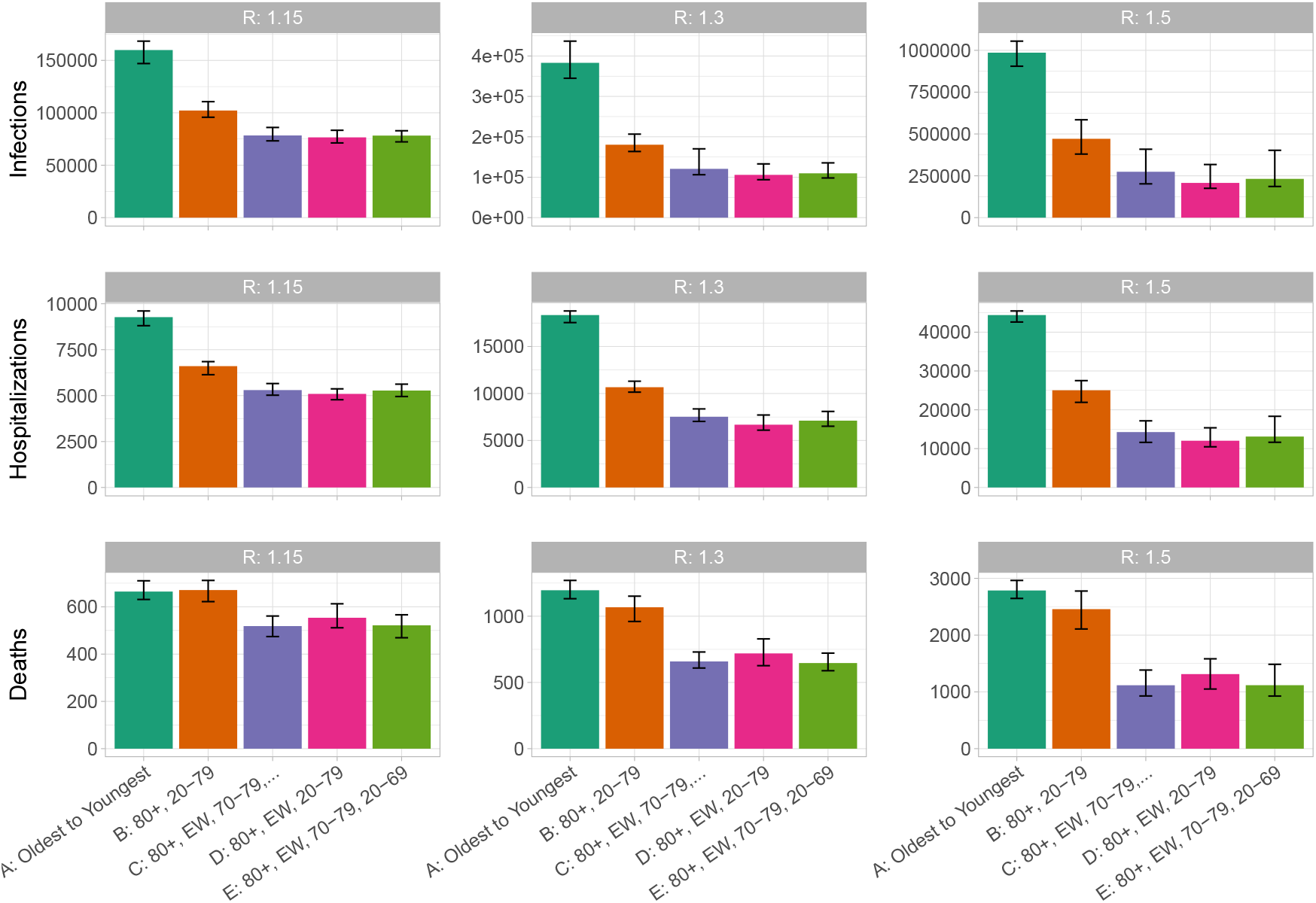
Sensitivity of results with respect to randomly resampling the contact matrix. Bars indicate the median value, with 5 and 95% quantiles shown as error bars. Fixed *α* = 0, *𝓋*_*e*_ = 0.6, *𝓋*_*p*_ = 0.9. For each simulation, perturb *C* by taking 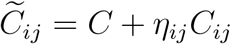, where *η* is a symmetric random matrix with each entry drawn from a normal distribution with mean 0 and standard deviation 0.3. We take *η*_*ij*_ = *η*_*ji*_ to preserve the contact structure of *C*.

**Figure S10:**
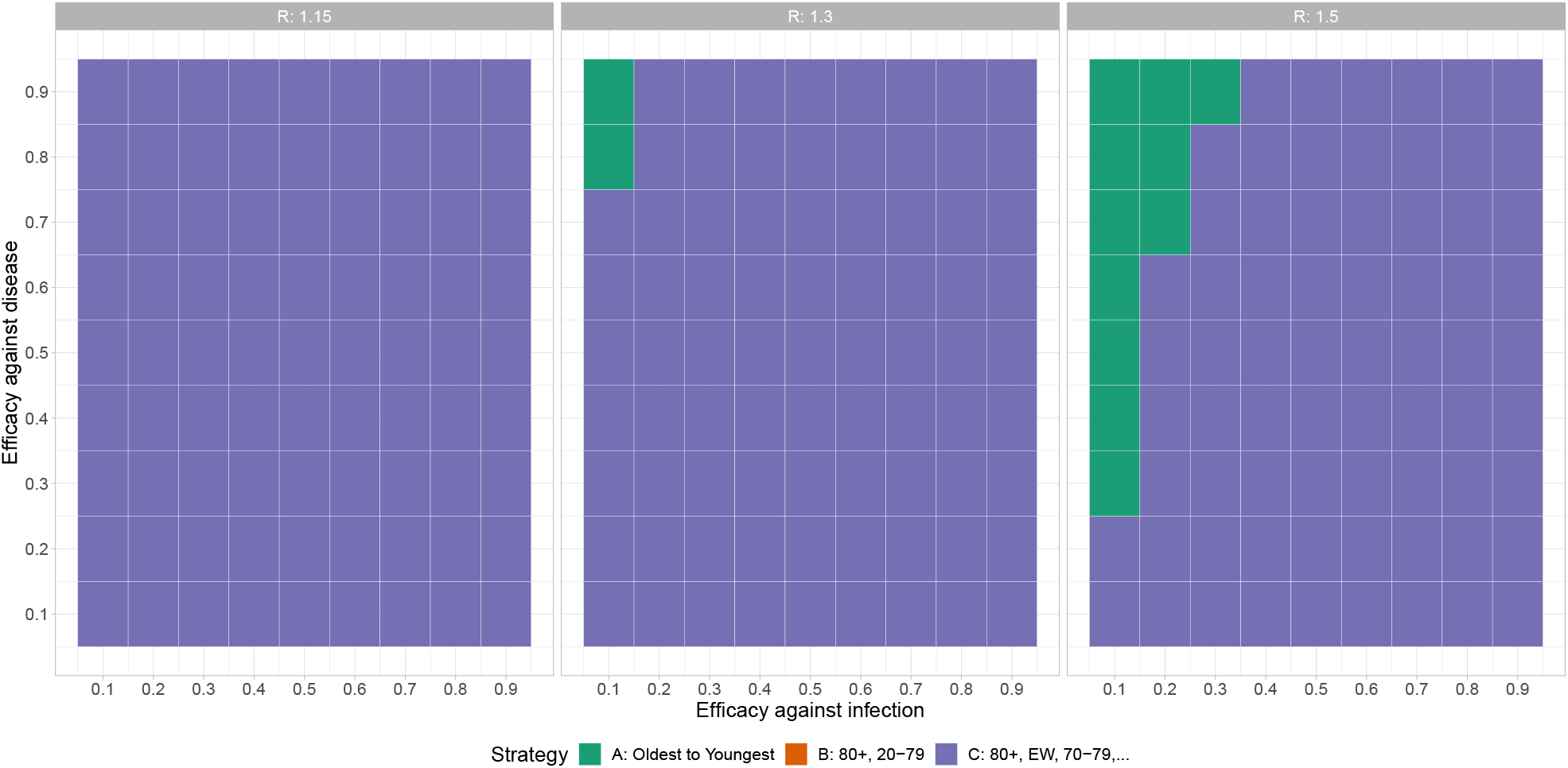
Optimal strategy in terms of minimizing deaths, as a function of *𝓋*_*e*_ and *𝓋*_*p*_. The rate of vaccination is fixed at 0.3 percent per day, and here we do not use a two-stage simulation. *R* is varied from 1.15 (left) to 1.3 (right). Note that other strategies which prioritize essential workers (Strategies D,E,F,G) often perform well also.

**Figure S11:**
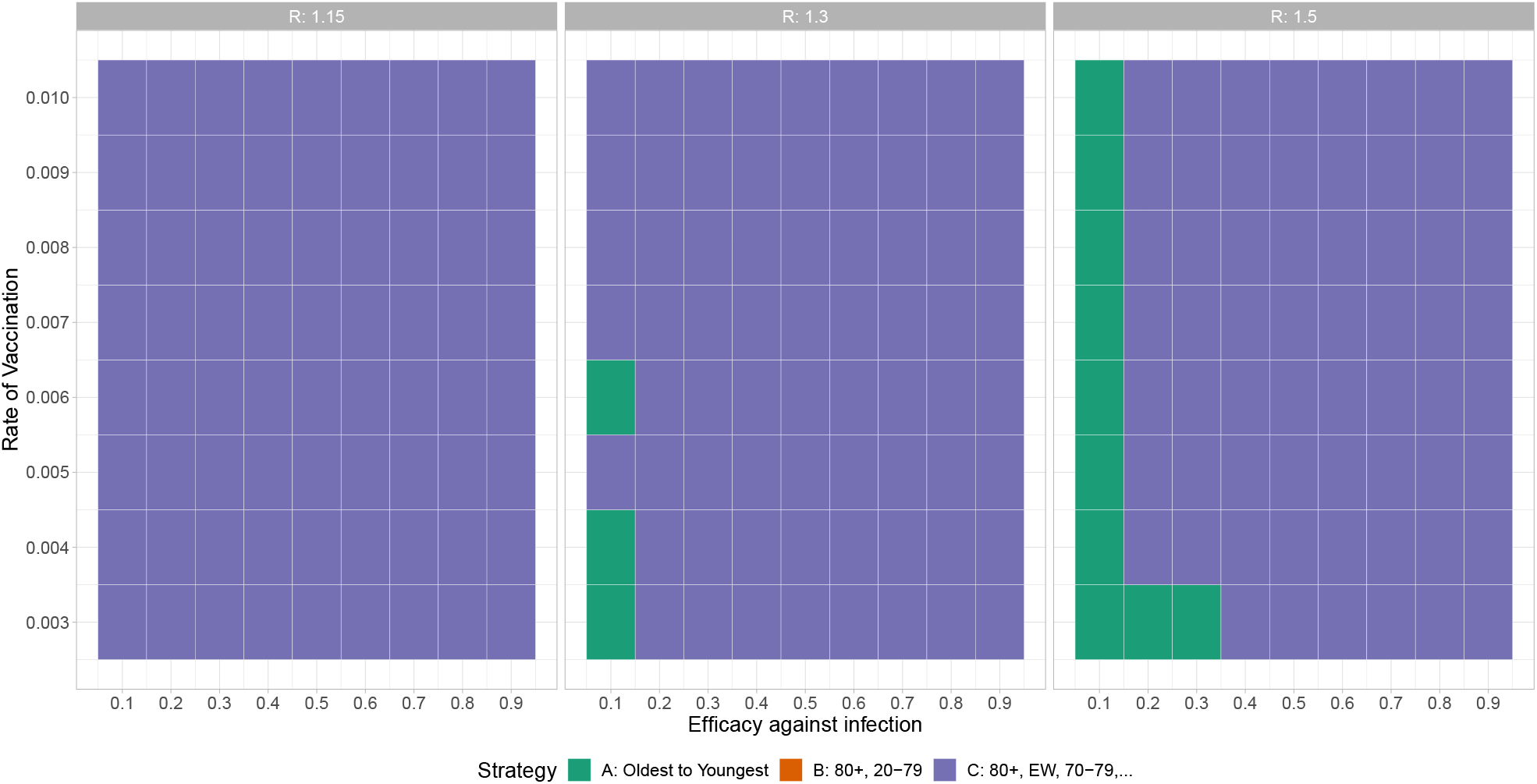
Optimal strategy in terms of minimizing deaths, as a function of *𝓋*_*e*_ and the rate of vaccination. Fixed *𝓋*_*p*_ = 0.9, and here we do not use a two-stage simulation. *R* is varied from 1.15 (left) to 1.3 (right). Note that other strategies which prioritize essential workers (Strategies D,E,F,G) often perform well also. Vaccination rate is varied from 0.3% of the population per day to 1% of the population per day.

## Footnotes

1 https://github.com/nmulberry/essential-workers-vaccine

## Notes

### Competing Interest Statement

The authors have declared no competing interest.

### Funding Statement

NM, PT, and CC were supported by Natural Science and Engineering Research Council (Canada) Discovery Grants (RGPIN-2019-06911 and RGPIN-2019-06624). CC receives funding from the Federal Government of Canada's Canada 150 Research Chair program and from Genome BC (COV-142). CM and EK were supported by an Alberta Health Partnership grant. No author received payment or services from any third party for any aspect of this work. Funders had no role in the study design, preparation or analysis.

### Author Declarations

No approval was required.

